# A Multiethnic Analysis of DNA Methylation Reveals Links Between Alzheimer’s Disease and Metabolic Comorbidities

**DOI:** 10.64898/2026.10.05.26364777

**Authors:** Benjamin Landon, Ann Abraham Daniel, Tamara Fischer, Nicole R. Phillips, Robert C. Barber, Austin W. Reynolds, The Health and Aging Brain Study: Health Disparities (HABS-HD) Study Team

**Author notes:** Corresponding Author: Austin W. Reynolds, Department of Microbiology, Immunology and Genetics, College of Biomedical and Translational Sciences, University of North Texas Health Science Center, Fort Worth TX, USA. HABS-HD MPIs: Sid E O’Bryant, Kristine Yaffe, Arthur Toga, Robert Rissman, & Leigh Johnson; and the HABS-HD Investigators: Meredith Braskie, Kevin King, James R Hall, Melissa Petersen, Raymond Palmer, Robert Barber, Yonggang Shi, Fan Zhang, Rajesh Nandy, Roderick McColl, David Mason, Bradley Christian, Nicole Phillips, Stephanie Large, Joe Lee, Badri Vardarajan, Monica Rivera Mindt, Amrita Cheema, Lisa Barnes, Mark Mapstone, Annie Cohen, Amy Kind, Ozioma Okonkwo, Raul Vintimilla, Zhengyang Zhou, Michael Donohue, Rema Raman, Matthew Borzage, Michelle Mielke, Beau Ances, Ganesh Babulal, Jorge Llibre-Guerra, Carl Hill and Rocky Vig.

## Abstract

**Background:** Alzheimer’s disease (AD) is the leading contributor to dementia burden, and the risk of developing it is impacted by a multitude of factors. Among these factors are metabolic conditions like type-2 diabetes (T2D) and metabolic syndrome (MetS), along with ethnicity, which interacts both with AD and the metabolic comorbidities. Previous studies have established associations between methylation patterns at specific CpG sites and risk for AD, T2D and MetS individually, but the underlying etiology that is responsible for interactions between metabolic conditions and AD risk remains unresolved.

**Objective:** We aimed to examine methylation patterns among Mexican American (MA) and Non-Hispanic White (NHW) individuals in the context of AD, alongside T2D and MetS, to identify CpG sites associated with disease-specific and interaction-dependent methylation effects related to those comorbidities.

**Methods:** DNA methylation data from 551 patients of MA or NHW ancestry who were enrolled in the Texas Alzheimer’s Research and Care Consortium (TARCC) were available for analysis. Participants were stratified into groups by AD/T2D status or AD/MetS score and Beta-values from the Illumina MethylationEPIC chip array were obtained for each patient. Differential methylation status was analyzed using the lumi and cate packages in R.

**Results:** Five CpG sites had statistically significant differential methylation patterns, including two (cg06322660 and cg25514677) which exhibited significant interactions between AD and T2D status or AD and MetS score respectively, with cg06322660 being significant in the NHW cohort and cg25514677 being significant in the MA cohort. The association at cg25514677 was replicated in an independent MA cohort.

**Conclusions:** The two sites associated with interactions between AD and a metabolic condition could explain the influence that those conditions have on the risk for AD and are prime candidates for future study.

## Introduction

Alzheimer’s Disease (AD) is the leading form of dementia worldwide, accounting for roughly two-thirds of all dementia cases [1]. This high degree of prevalence among the aging global population, combined with the enormous socioeconomic burden of the disease, has placed it high on the priority list of researchers around the world [2]. However, despite extensive effort[3] and abundant hypotheses [4], a definitive understanding of the underlying pathology of AD remains elusive and the vast majority of treatments that have been tested fail to translate during clinical trials [3]. At the clinical level, the strongest evidence we have to explain AD pathology comes from observing the highly conserved risk factors associated with the disease [5–9].

In addition to age and education level, a wide range of factors are reliably observed to contribute to AD risk, including metabolic conditions such as metabolic syndrome (MetS) and type-2 diabetes (T2D) [10–14]. There is some evidence that insulin resistance plays a role outside of its typical effects on glucose uptake, and metabolic syndrome could be involved through a variety of inflammatory and circulatory mechanisms [12,13,15–17]. While these associations suggest that interrelated pathways contribute to shared risk between metabolic diseases and AD, we still lack a clear understanding of which genes may be contributing to this interaction.

Adding to the complexity of the risk model is the fact that both AD and metabolic conditions disproportionately impact certain populations. For instance, MetS and T2D are roughly twice as common among Mexican American (MA) individuals, who are also 1.5 times more likely than non-Hispanic White (NHW) individuals to develop AD [2,18]. In addition, genetic risk factors for AD also vary substantially by ethnicity [19–22]. While the *APOE* ε4 allele confers the largest amount of risk of any known AD associated loci [2,23], its effect size varies across genetic an-cestries [24,25]. Several large-scale genome wide association studies (GWAS) have been conducted for AD [6,26–28] to identify further genetic risk factors. Unfortunately, the cumulative effects of the risk loci with the largest effect sizes still only account for a small portion of the heritability of AD risk, making the current generation of polygenic risk scores unsuitable for accurately predicting development of the disease [7–9,29,30].

DNA methylation patterns offer a potentially useful tool to explain some of the additional variation not captured by the genetic sequence alone and may account for some of the unexplained risk. Previous studies have identified specific DNA methylation patterns associated with AD [31–34] and MetS separately [35], but little work has been done on shared methylation patterns between the two conditions. Here we extend the approach used in a previous study from our team, which examined methylation related to AD diagnosis between ethnicities [31], to include an epigenome-wide study of metabolic syndrome in the context of AD and further refine our understanding of the interaction between these two conditions.

### Methodology

#### Study design

Study participants (N=551) came from the Texas Alzheimer’s Research and Care Consortium (TARCC), which aggregated blood samples, cognitive, clinical, molecular and demographic data for a case/control cohort study of AD. Demographic data for the sample are visualized in Table 1. Patients with a consensus diagnosis of either AD or MCI (mild cognitive impairment) were categorized as having cognitive impairment (CI), while those without an AD or MCI diagnosis were considered cognitively unimpaired (CU). Included in the patient clinical data was information on diabetic, hypertension, and hypercholesterolemia status. The T2D analysis focused on diabetic status in conjunction with cognitive impairment (CI). Due to limitations in the available clinical data, our MetS score was computed in a non-traditional manner. Typically a patient would be classified as having metabolic syndrome if they have 3 out of the 5 diagnostic conditions (diabetes, hypertension, hypercholesterolemia, elevated fasting glucose, and obesity as determined by waist circumference)[36]. Because our data lacked relevant measures for fasting glucose and waist circumference, we instead used a system that summed the number of conditions they were positive for among the three we had available, giving us a score range of 0 to 3. While this technique of scoring has not been utilized directly before, there is evidence to show that MetS associated risk is cumulative from its individual components [37,38]. In addition, participant data was pulled from the Health and Aging Brain Study: Health Disparities (HABS-HD) to perform replication analysis on the hits we found in the TARCC cohort, utilizing the same criteria and diagnostic measures.

**Table 1:** Demographics Data Two-sample t-tests with assumption of unequal variance were used to examine significant differences between unimpaired (CU) and impaired (CI) individuals on the basis of age (years), education level (years), Mini-Mental State Exam (MMSE) scores, Clinical Dementia Rating (CDR) sum scores, and average MetS scores. Chi-Square test was used to examine differences in T2D positivity rates between CU and CI groups and male/female group proportions.

| Non-Hispanic Whites n = 252 |  |  |  |
| --- | --- | --- | --- |
|  | CU | CI (AD/MCI) | p |
| n | 143 | 109 | - |
| Sex (F/M) | 72/71 | 51/58 | ns |
| Age (y) | 71.1 ± 7.4 | 72.1 ± 4.5 | ns |
| Education (y) | 16.0 ± 2.6 | 14.9 ± 2.7 | <0.01 |
| T2D (positive:negative) | 47:96 | 38:71 | ns |
| MetS (mean + sd) | 2.489 + 1.093 | 2.688 + 0.930 | ns |
| MMSE | 29.2 ± 1.0 | 23.9 ± 5.1 | <0.01 |
| CDR Sum | 0.0 ± 0.1 | 3.8 ± 3.2 | <0.01 |
| Mexican Americans n = 299 |  |  |  |
|  | CU | CI (AD/MCI) | p |
| n | 177 | 122 | - |
| Sex (F/M) | 101/76 | 65/57 | ns |
| Age (y) | 67.6 ± 5.6 | 70.7 ± 7.8 | <0.01 |
| Education (y) | 11.0 ± 4.4 | 9.1 ± 5.3 | <0.01 |
| T2D (positive:negative) | 76:101 | 50:72 | ns |
| MetS (mean + sd) | 2.672 + 1.014 | 2.729 + 0.909 | ns |
| MMSE | 28.1 ± 2.0 | 23.6 ± 5.0 | <0.01 |
| CDR Sum | 0.0 ± 0.1 | 2.4 ± 3.0 | <0.01 |

#### Analysis of methylation data

Beta values, representing the likelihood of a site being methylated between 0 and 1, were acquired from the Infinium^©^ MethylationEPIC BeadChip array (Illumina). These values were normalized using the Beta Mixture Quantile dilation (BMIQ) method [39], then converted into M values using the lumi R package before using the cate R package alongside patient covariate data to perform differential methylation analysis [40–44]. Models controlled for age, sex, estimated cell proportions, and genetic ancestry through PCA eigenvectors 1 and 2, while looking for interactions between cognitive status and either T2D status or MetS score. In addition to these covariates, we also estimated the number of missing technical factors from the dataset to reduce genomic inflation and prevent false-positive results using cate’s “est.confounder.num” function. In the T2D analysis, 10 and 6 were estimated to be the optimal latent factor values for the MA and NHW groups respectively, meaning cate will estimate and account for that number of unobserved covariates when performing each analysis. When utilizing MetS, which incorporates T2D as a measure, these estimated optimal values were 11 and 7 respectively. Relevant QQ-plots for each cate model are available in the supplemental materials (supplemental figures 1-20). The model equations used in cate can be seen below (formulae 1-4). These models were run independently on both the MA and NHW groups. P-values from the cate outputs were adjusted using the Benjamini-Hochberg False Discovery Rate (FDR) correction (alpha = 0.05). Significant sites following correction for multiple testing were paired with their genome location, and relevant genes determined either by known CpG hits in the MethylationEPIC probe features data, or by manual examination of the site genome region on the UCSC Genome Browser (GRCh37/hg19) [45].

Formula 1 (MA/T2D):

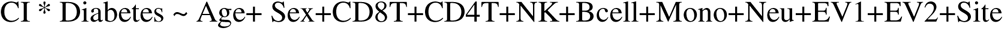

Formula 2 (NHW/T2D):

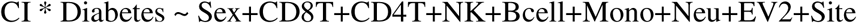

Formula 3 (MA/MetS):

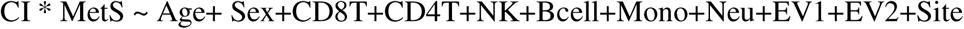

Formula 4 (NHW/MetS):

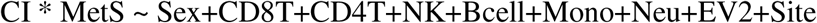

## Results

We conducted epigenome-wide association studies (EWAS) on 551 previously sampled participants from the TARCC to investigate the relationship between AD and cardiometabolic diseases as mediated through DNA methylation [31]. Our analysis was split between MA (N=299) and NHW (N=252) populations on the basis of both cognitive status and metabolic co-morbidities (Table 1). The MA cohort had an average age of 67.6 and 70.7 years for CU and CI respectively, which differed significantly (p < 0.01), while the NHW cohort had 71.1 and 72.1 years, respectively, with no significant difference between them. Similarly, the male to female proportions between CU and CI did not differ significantly, nor did the average MetS scores or T2D positivity rates, for either ethnicity. Incorporating the metabolic factors alongside the cognitive diagnosis enabled us to isolate which sites could be responsible for each factor’s contribution to dementia risk, and significant sites were found for each.

### Significant differentially methylated sites in MAs

We identified a single significant CpG site (cg25514677) among MAs that was associated with the interaction between MetS and CI (Figure 1B), following correction for multiple testing. cg25514677 is closely located to a novel RNA encoding gene, which is itself associated with the genes *FOSL2* and *RBKS*, due to their shared ability to be targeted by similar gene enhancers [45–50]. The association with this site was a negative, third-degree interaction effect, indicating a complex pattern of exacerbated hypomethylation driven by the presence of metabolic syndrome in conjunction with CI (p = 0.001).

**Figure 1:**
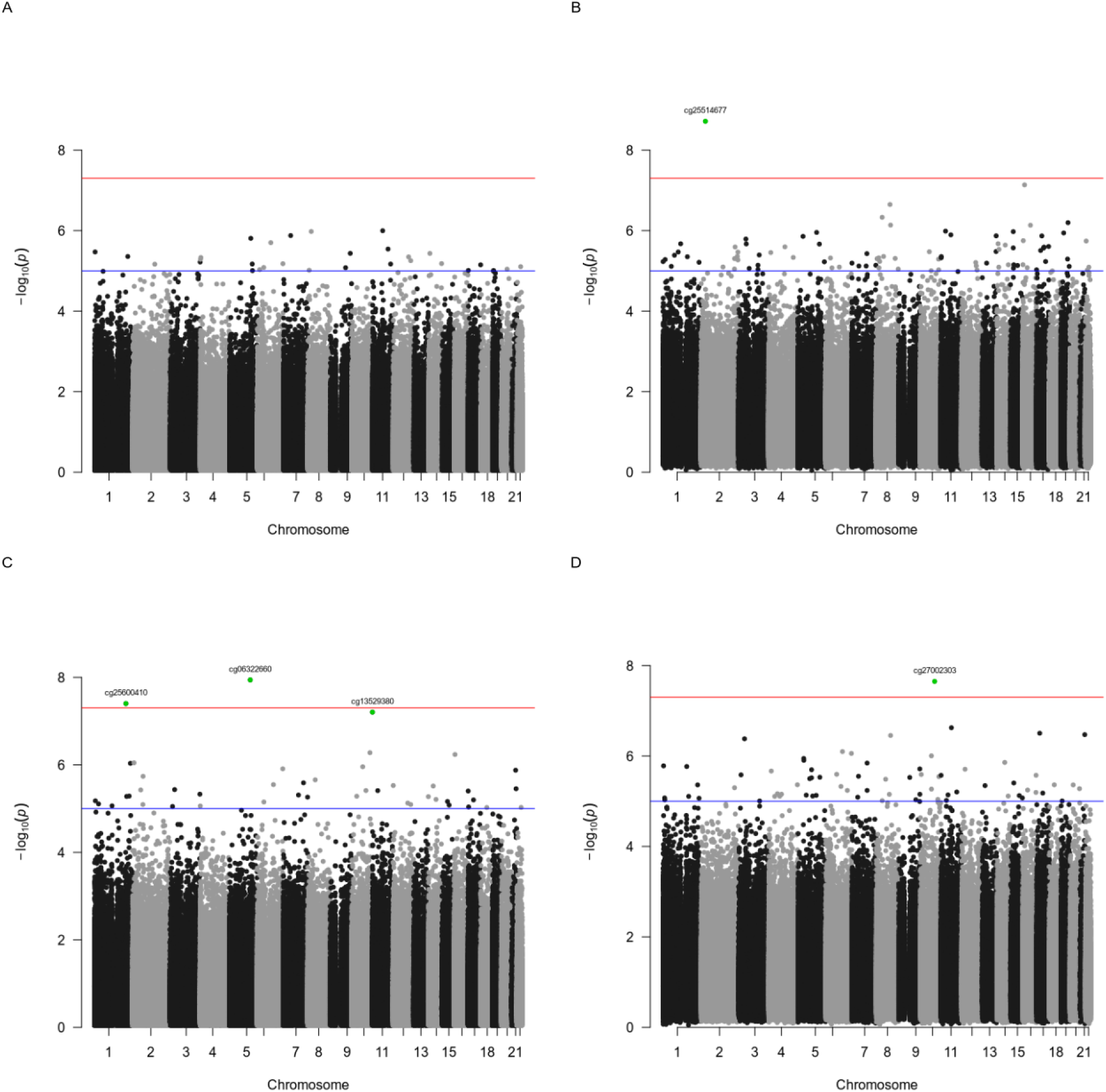
Manhattan Plots of Diabetes and MetS Associated Methylation Sites Manhattan plots mapping significance levels of differential methylation at CpG sites across the genome for A) MA + T2D. B) MA + MetS. C) NHW + T2D. D) NHW + MetS. The red line indicates a raw p-value threshold of 5e-08, while the blue line indicates a cutoff of 1e-05. Sites that were significant after adjustment (FDR p-value < 0.05) are highlighted in green and labeled.

### Significant differentially methylated sites in NHWs

In our EWAS of the NHW cohort, we identified three significant sites associated with the interaction of T2D and CI (Figure 1C). Among the T2D-associated sites, diabetes status drove a hypomethylation effect on cg25600410 (p = 0.029), located in the *MFSD4* gene. CI similarly showed a hypomethylation effect on cg13529380 (p=0.045), located in a novel transcript gene with anti-sense regions to *NLRP6* and *ATHL1* [45,51]. Finally, we found an interaction effect between T2D and CI on cg06322660 (p=0.008), located in the *CTB-1I21.1* gene, driving a hypermethylation effect at the site [45]. We also identified a single significant site (cg27002303), associated with CI when controlling for MetS (Figure 1D). This site is located in a novel RNA transcript gene anti-sense to the *HECTD2* gene, a component of the ubiquitin ligase system [45,52,53]. At this site, we saw a hypermethylation effect associated with CI (p = 0.016).

#### Replication in separate cohort

We attempted to replicate our significant findings using data from an additional cohort of 493 Mexican American and 562 Non-Hispanic White patients from the Health and Aging Brain Study (HABS-HD). Cohort case and control counts are given in Table 3. All methods for the analysis were the same, and all variables used to analyze the TARCC cohort were available and used for the HABS-HD cohort, with the exception of the PCA derived eigenvectors EV1 and EV2. However, the cate function for estimating the number of confounding factors suggested that these were not necessary to capture the variability within the HABS-HD cohort. We are only able to replicate the association at cg25514677 with an interaction between CI and MetS (p = 0.004) in the HABS-HD cohort (Table 2). The effect size of both associations are in the same direction, but slightly lower in the HABS-HD replication (*b* = −0.012).

**Table 2:**
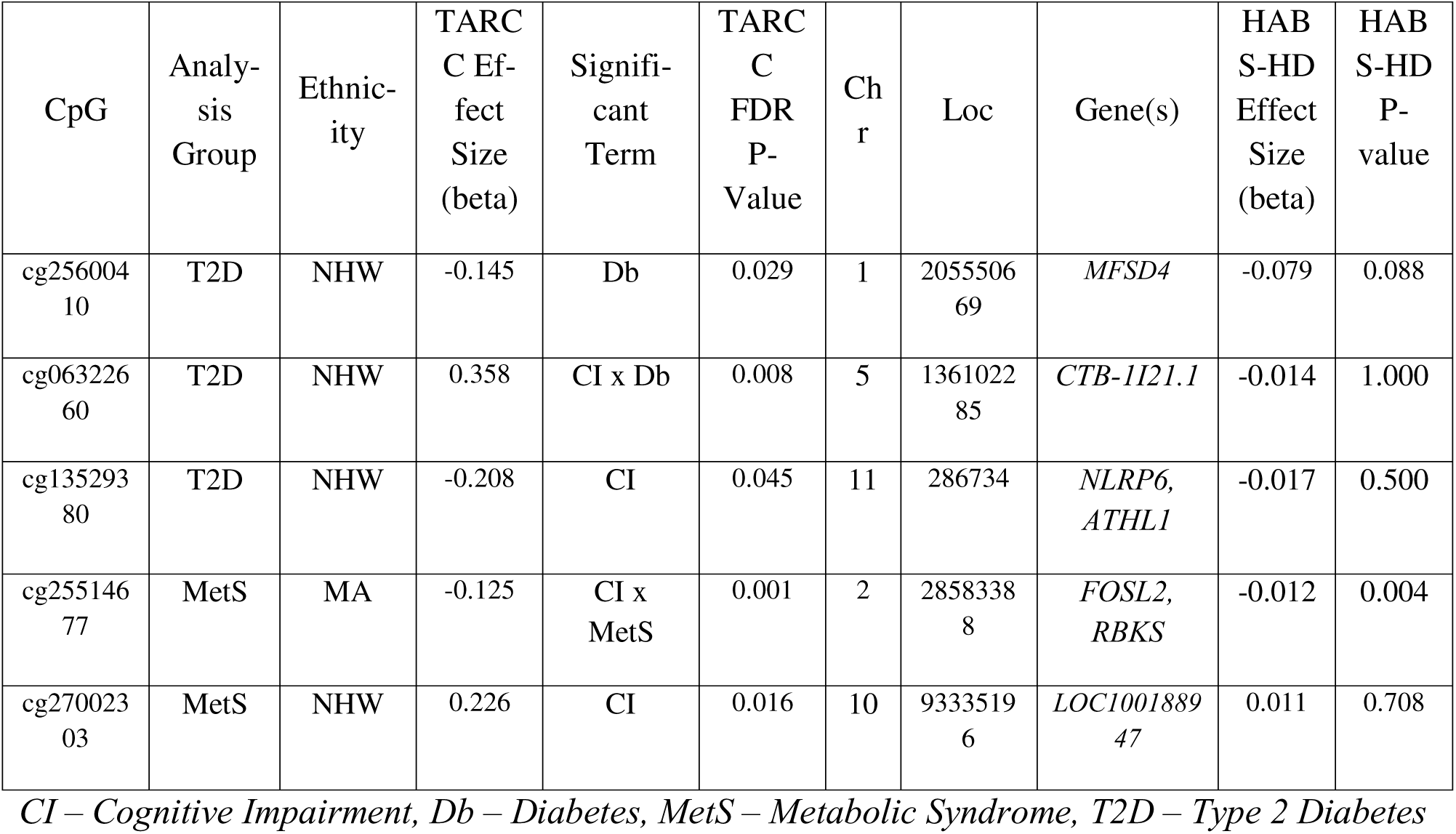
Significant CpG Sites and Interacting Genes.

**Table 3.** HABS-HD Case/Control Data - Sample Sizes.

| **Ethnicity** | **Gender** | **Cognitive Status** |  |
| --- | --- | --- | --- |
|  |  | **CU** | **CI (AD/MCI)** |
| White | Male | 184 | 48 |
|  | Female | 296 | 34 |
| Hispanic | Male | 122 | 39 |
|  | Female | 273 | 59 |

*CI - Cognitively Impaired, CU - Cognitively Unimpaired*

## Discussion

We analyzed the genome-wide methylation patterns from a cohort of age and sex-matched MA and NHW individuals, looking for differentially methylated sites linked to CI, T2D, and MetS score and identified five sites significantly associated with one or more of these conditions. Previous studies have identified numerous significant sites related to CI [54], and implicated methylation status in the risk for metabolic conditions like T2D [55]. Similarly, this study has found sites that correspond to risk for each condition independently. However, some of these sites are associated with an interaction between CI and one of the metabolic conditions, which could help explain the increases in AD risk being driven by MetS and T2D.

### Cognitive Impairment associated sites

Two CpG sites (cg13529380 and cg27002303) were found to be significantly associated with CI when controlling for MetS score or T2D, both of which were also detected in the previous study by Abraham Daniel *et al* [31]. The cg13529380 site falls within a long non-coding RNA [45,51]. This could potentially be a pri-miRNA, given its anti-sense sequences to the *NLRP6* and *ATHL1* genes [51]. Pri-miRNA are the initial, long transcripts that eventually get processed into micro RNA (miRNA) in the canonical biogenesis pathway [56]. These miRNA are known primarily for their regulatory activity, exerted by way of RNA interference or silencing, and alterations in the expression patterns of these miRNA have been observed in AD [57,58]. Both *NLRP6* and *ATHL1* are related to pathways implicated in AD, with *NLRP6* interacting with nuclear factor kappa B (NF-ΚB) and *ATHL1* relating to dysfunctional glucose metabolism [59,60]. That is made more interesting by the fact that cg13529380 is significantly associated with CI and not T2D status or MetS score. This could imply that the downstream process being controlled by this site is separate from the characteristic insulin dysfunction seen in those comorbidities.

The cg27002303 site is located in a region encoding an anti-sense RNA for the *HECTD2* gene, a component of E3 ubiquitin ligase 2 [45,52,53]. *HECTD2* is also associated with susceptibility to prion diseases [61,62]. This, in conjunction with the proteolytic ubiquitin association, provides a basis by which the CpG site could be interacting with AD through expression changes of proteasome components, given the established proteostasis disruptions associated with the disease [63–66]. Increased methylation at this site could reduce the transcription of that anti-sense RNA, and thus potentially increase expression of *HECTD2* by preventing the potential RNA interference.

### Diabetes-associated sites

There was a single site associated specifically with T2D when controlling for cognitive status, cg25600410, located in the *MFSD4* gene, which regulates membrane glucose transporters [67]. Issues with glucose metabolism can affect the brain in dementia, primarily through resistance to insulin’s effects that do not relate to glucose transport across cellular membranes [15,16,68].

This is because the brain’s glucose transporters are not insulin dependent [16]. It is possible that *MFSD4* could interact with the brain’s insulin-independent GLUT3 transporters to disrupt metabolism and contribute to AD risk, but this transcript appears to have biased expression in the kidneys and stomach, rather than the brain [67]. This suggests that this site is more likely related to diabetic processes than dementia processes directly.

### Sites associated with interactions between Cognitive Impairment and metabolic phenotypes

Two sites were found to be significantly associated with an interaction between cognitive impairment and a metabolic phenotype in our dataset. The site cg06322660 was associated with an interaction effect between T2D and CI in the NHW cohort. This site is closely positioned to the gene *CTB-1I21.1*, which exhibits biased expression in the brain, [69], specifically in the amygdala and hypothalamus [45,70], making this site of key interest for potential diabetes incurred risk of AD [71–74]. Furthermore, while no sites were found to be significantly associated with MetS alone in our analysis, cg25514677 was found to be significantly associated with interaction effect between MetS score and CI. The negative interaction effect suggests that methylation at this site was lower in cognitively impaired individuals, and this was exacerbated by metabolic dysfunction. The negative interaction effect at cg25514677 is also the only significant finding from our results that is replicated in an independent methylation dataset from the HABS-HD cohort. The presence of this effect in both cohorts further strengthens the evidence for its contribution to the MetS associated risk of dementia, and warrants further investigation into the site.

This cg25514677 site itself is located between two genes that are both affected by the same enhancing transcription factors, meaning altered methylation at this site could affect accessibility of those nearby genes for expression [45]. One of them, *FOSL2*, is itself a transcription factor for *AP-1*, which is a more general transcription factor involved in regulating many genes, including not only components of the ubiquitin ligase proteasome, but also amyloid-β precursor protein (AβPP) [75–77]. The resulting potential for increased AβPP expression is an interesting facet of these findings and could offer a potential explanation of how AβPP levels could increase and drive production of the characteristic Aβ_1-42_ in AD patients.

#### Limitations

This study was not without limitations. Firstly, the majority of our hits came from the NHW population. The one exception was cg25514677 which had the interaction effect between CI and MetS score in MAs. This is a surprising result, as we would expect more hits from the MA population, given their increased risk for both AD and the common metabolic co-morbidities we examined [78–80]. This could have been driven by our lack of all 5 traditional diagnostic conditions that define MetS. In a study that includes all of the measures available, it’s possible that more patients would fall under the diagnostic category of MetS, which could increase the number or strength of significant hits.

While this analysis was run on both MA and NHW individuals, these models were run independently, preventing us from looking directly at effects driven by ethnicity. However, this was not the primary focus of the study. Similarly, while we had sex included as a covariate in our models, it was not examined as a primary factor, despite its strong effects on AD risk. This is again due to the more narrow scope of the present study, but could be addressed in a future study to determine if any of these sites are being altered in a sex-specific fashion.

Additionally, while the overall number of samples in both MA and NHW models were relatively large, when we break down the group sizes involved in the interaction effects between CI and either T2D or MetS scores, we have between 30 and 50 participants per group, which suggests we may not be adequately powered to detect biologically relevant sites of lower effect sizes from both the MA and NHW populations.

Ultimately, the inclusion of MetS scores and T2D status as controlling factors in existing methylation data has helped us identify additional sites of interest compared to previous studies looking at cognitive impairment alone. This study design to categorize a site as relevant to AD, a metabolic comorbidity, or to both, should guide future research and allow us to better study how these comorbidities are influencing disease risk, as well as determine which sites are the most relevant for disease prediction. In both the MA and NHW populations, we identified sites where methylation was either associated with an interaction between cognitive status and a metabolic condition, or with a metabolic condition alone but targeting a gene relevant to dementia. This suggests that while the specific sites may vary across populations, they can impact the same biological pathways, contributing to the observed complex interactions between dementia and metabolic conditions. The site cg25514677, which was associated with a negative interaction between CI and MetS is of particular interest, given its replication in an independent cohort. Future studies will allow us to continue refining our understanding of these sites, and the degree of risk each confers for their associated diseases.

## Supporting information

Supplemental Figures - QQ Plots

## Data Availability

Data available upon request to the authors

## Acknowledgements

This work was supported by materials from the Texas Alzheimer’s Research and Care Consortium (TARCC).

## Statements and Declarations

### Ethical considerations

Not applicable

### Consent to participate

Not applicable

### Consent for publication

Not applicable

## Key

CI - Cognitive Impairment

MetS - Metabolic Syndrome Score

CD8T - Estimated CD8 T-cell proportion

CD4T - Estimated CD4 T-Cell proportion

NK - Estimated Natural Killer cell proportion

Bcell - Estimated B-Cell proportion

Mono - Estimated Monocyte proportion

Neu - Estimated Neuron proportion

EV1/EV2 - Eigenvectors 1 and 2 capturing genetic background

Site - TARCC recruitment site where patient data was collected.

