## Supplemental Figures - QQ Plots for "A Multiethnic Analysis of DNA Methylation Reveals Links Between Alzheimer’s Disease and Metabolic Comorbidities"

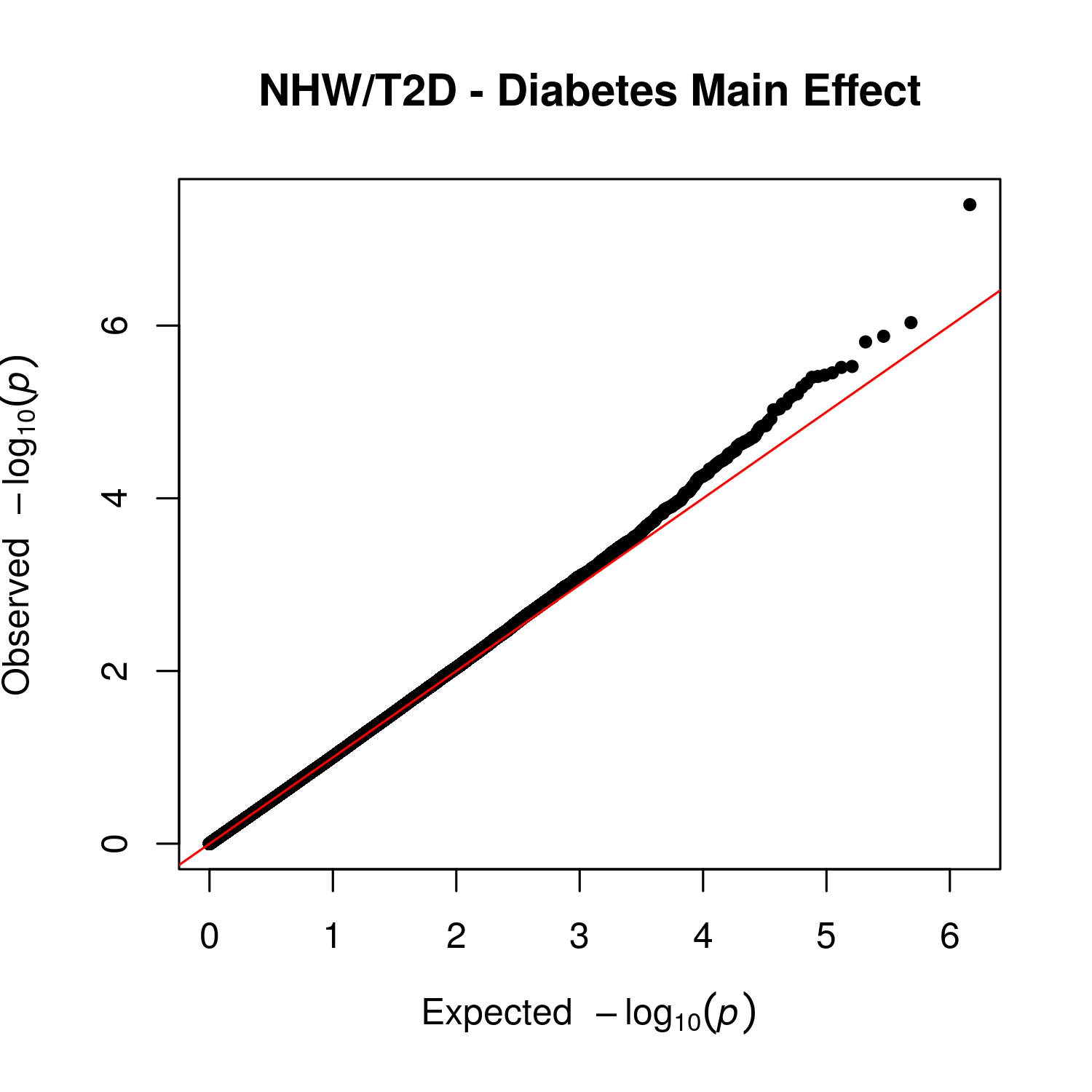

Supplemental Figure 1 – QQ Plot for Main Effect of Diabetes in Non-Hispanic White, Type 2 Diabetes Group

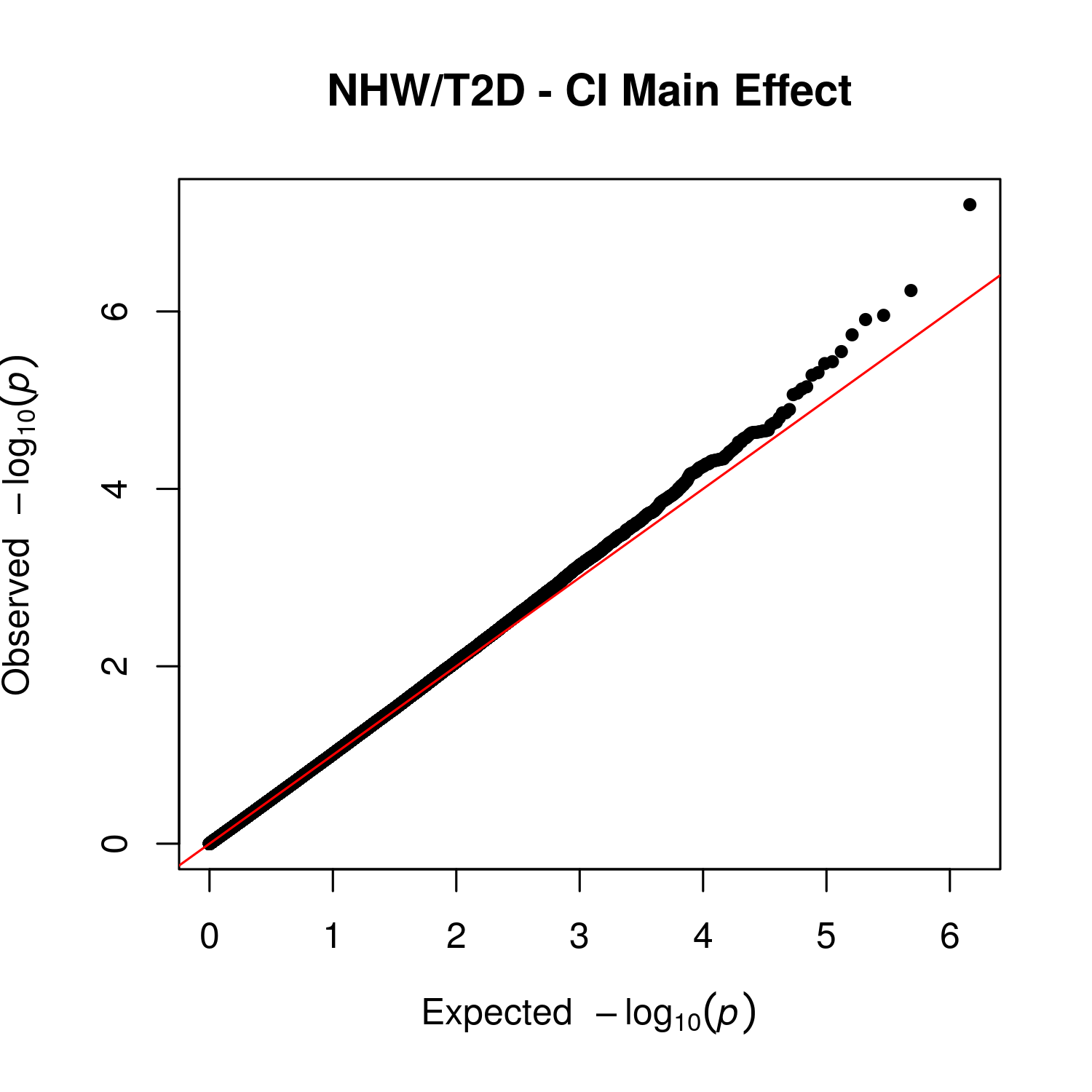

Supplemental Figure 2 – QQ Plot for Main Effect of Cognitive Impairment in Non-Hispanic White, Type 2 Diabetes Group

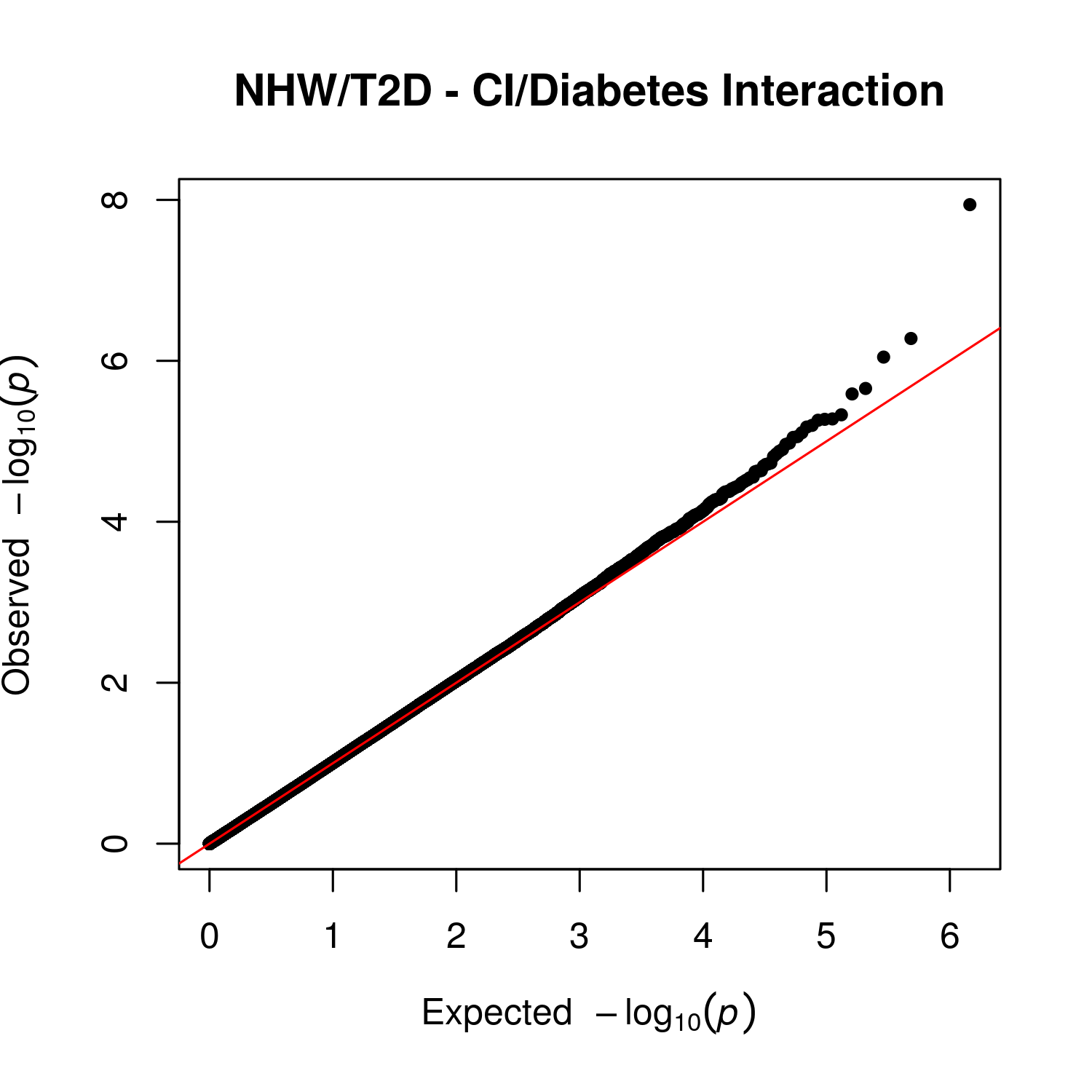

Supplemental Figure 3 – QQ Plot for Interaction Effect between Cognitive Impairment and Diabetes in Non-Hispanic White, Type 2 Diabetes Group

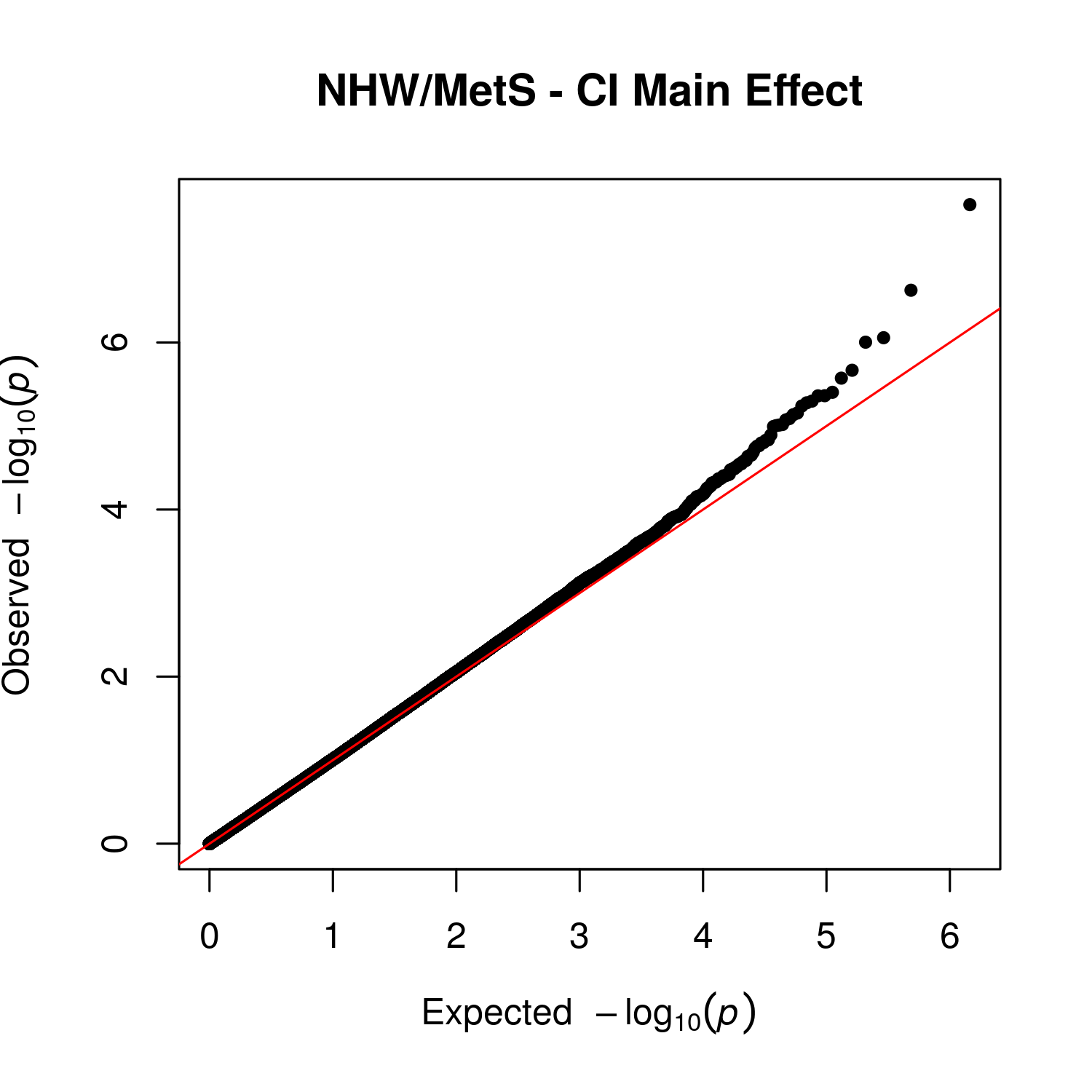

Supplemental Figure 4 – QQ Plot for Main Effect of Cognitive Impairment in Non-Hispanic White, Metabolic Syndrome Group

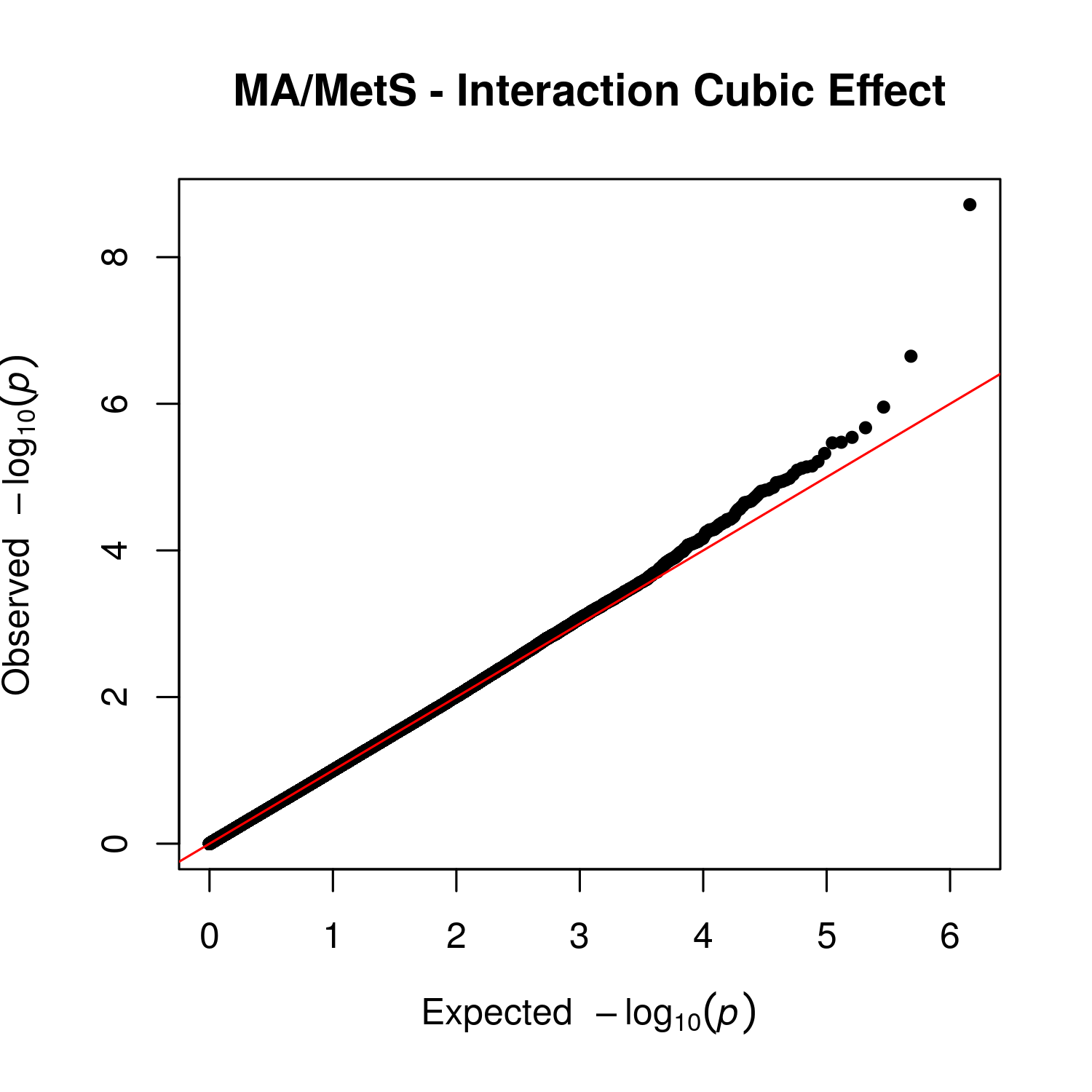

Supplemental Figure 5 – QQ Plot for Interaction Effect between Cognitive Impairment and MetS Score in Mexican American, Metabolic Syndrome Group

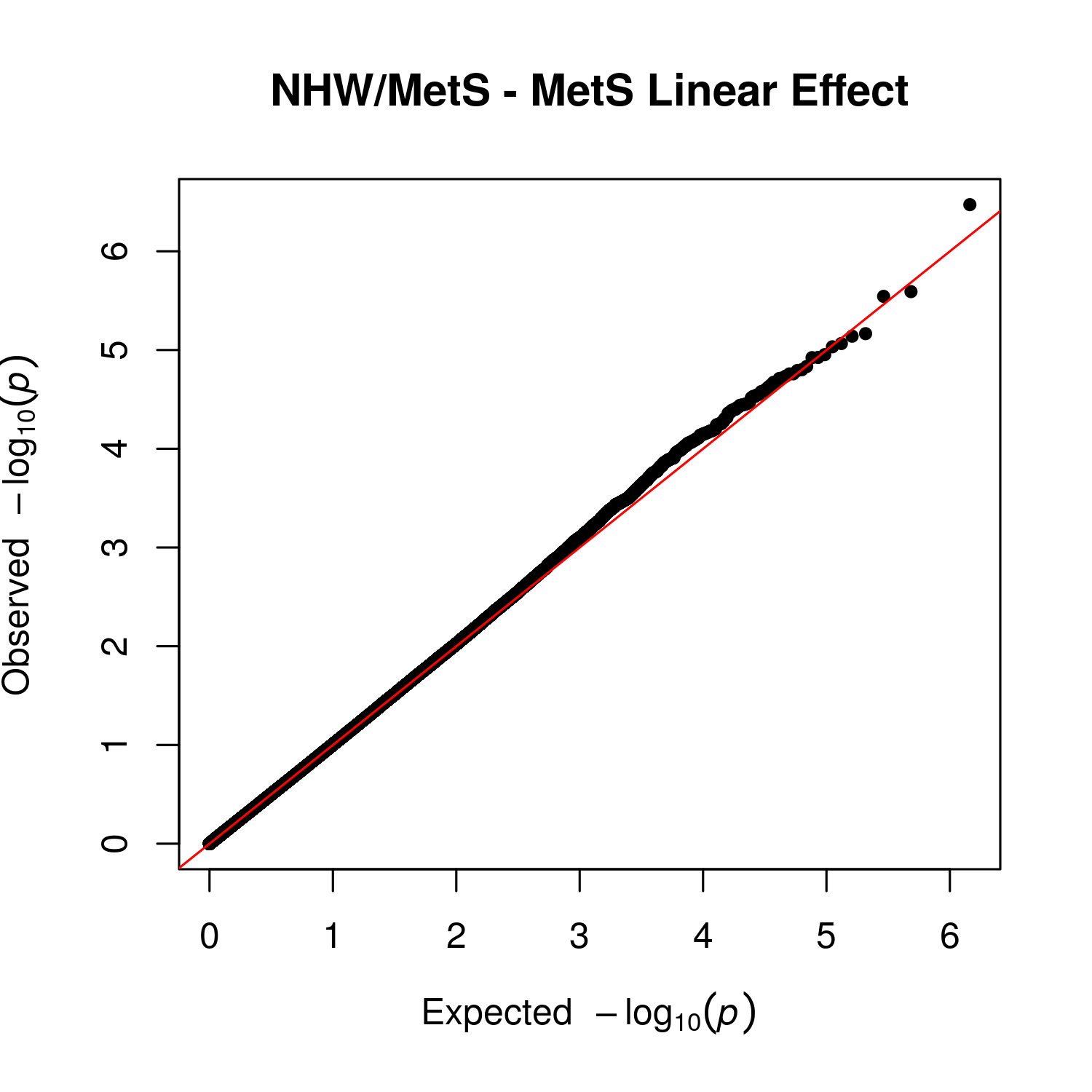

Supplemental Figure 6 – QQ Plot for Linear Main Effect of MetS Score in Non-Hispanic White, Metabolic Syndrome Group

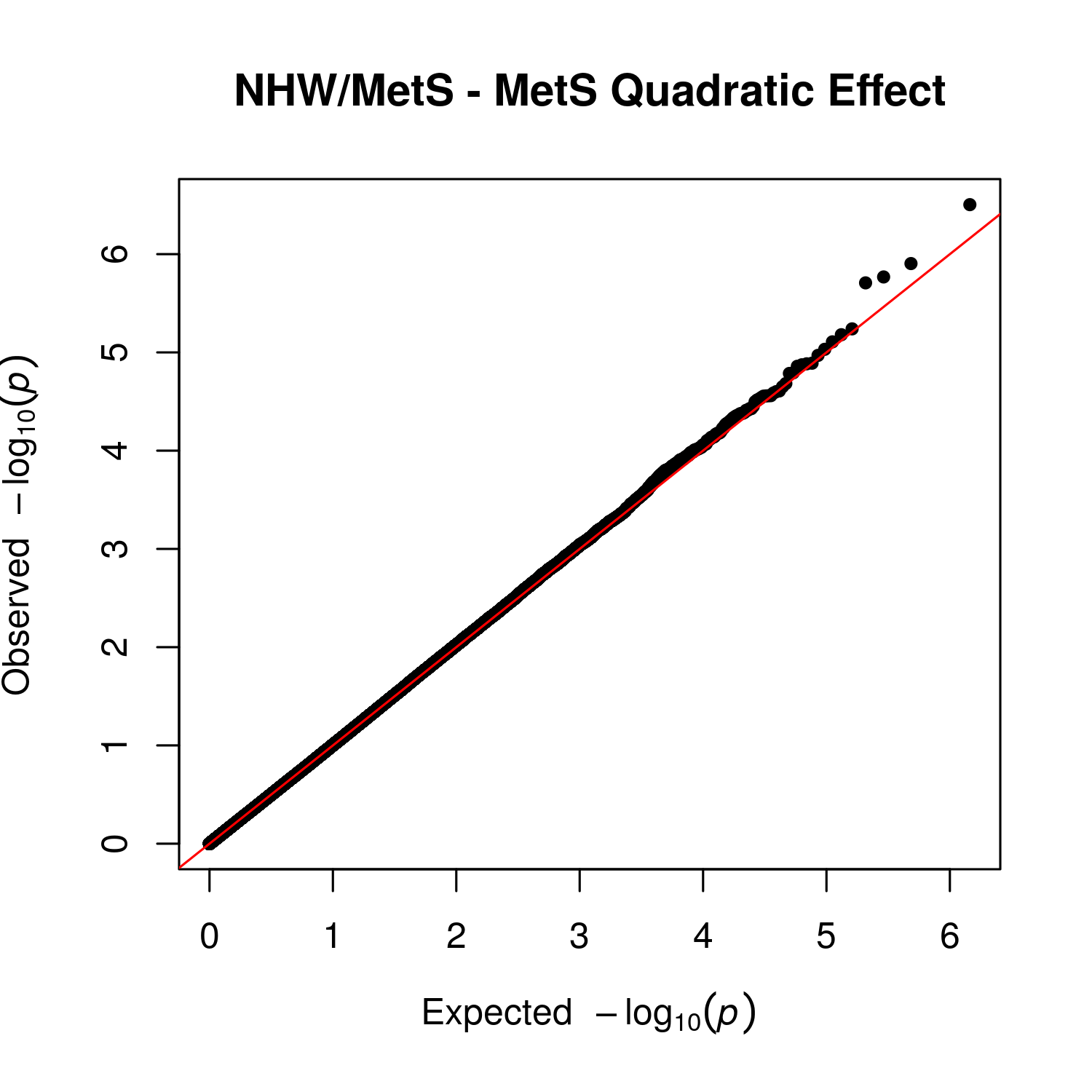

Supplemental Figure 7 – QQ Plot for Quadratic Main Effect of MetS Score in Non-Hispanic White, Metabolic Syndrome Group

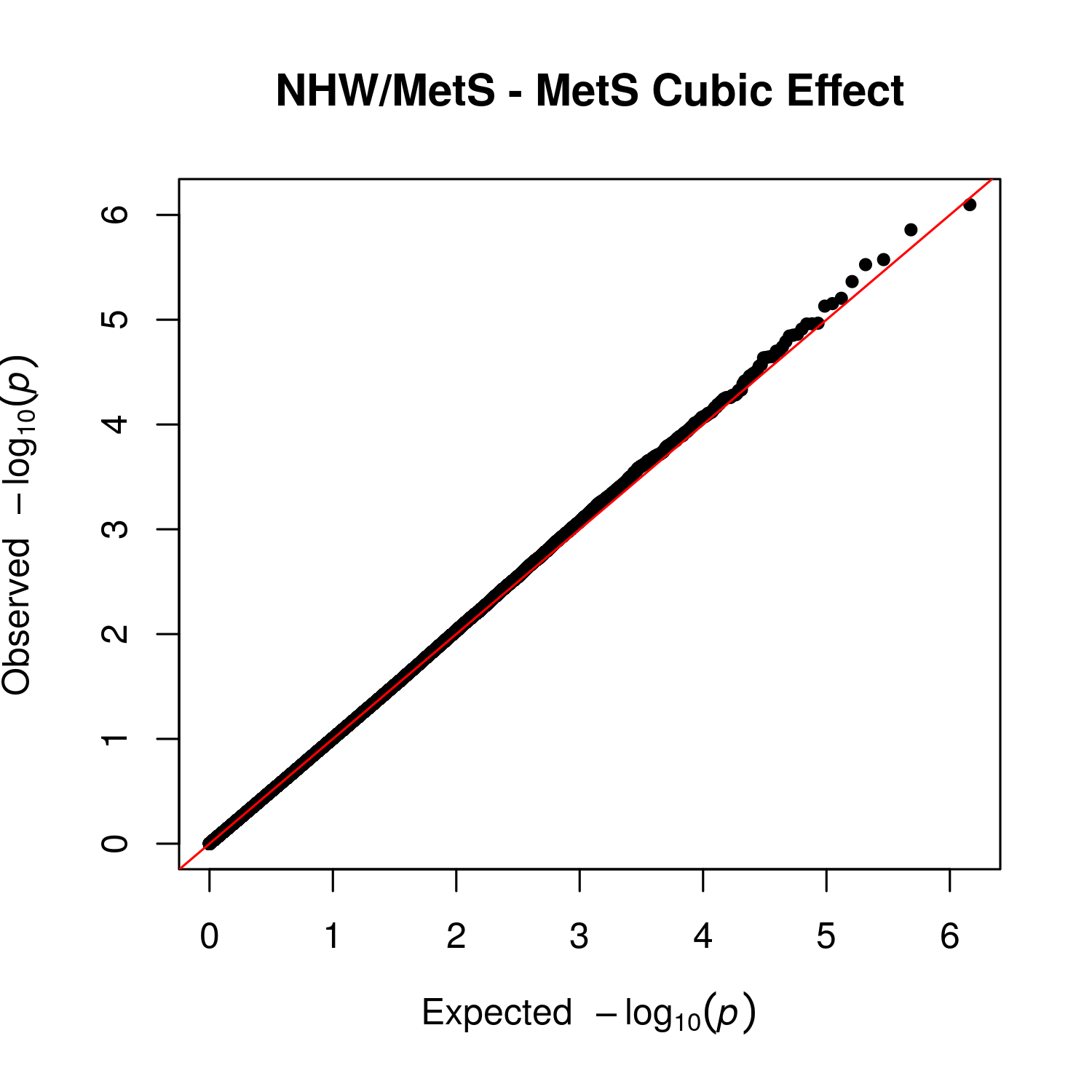

Supplemental Figure 8 – QQ Plot for Cubic Main Effect of MetS Score in Non-Hispanic White, Metabolic Syndrome Group

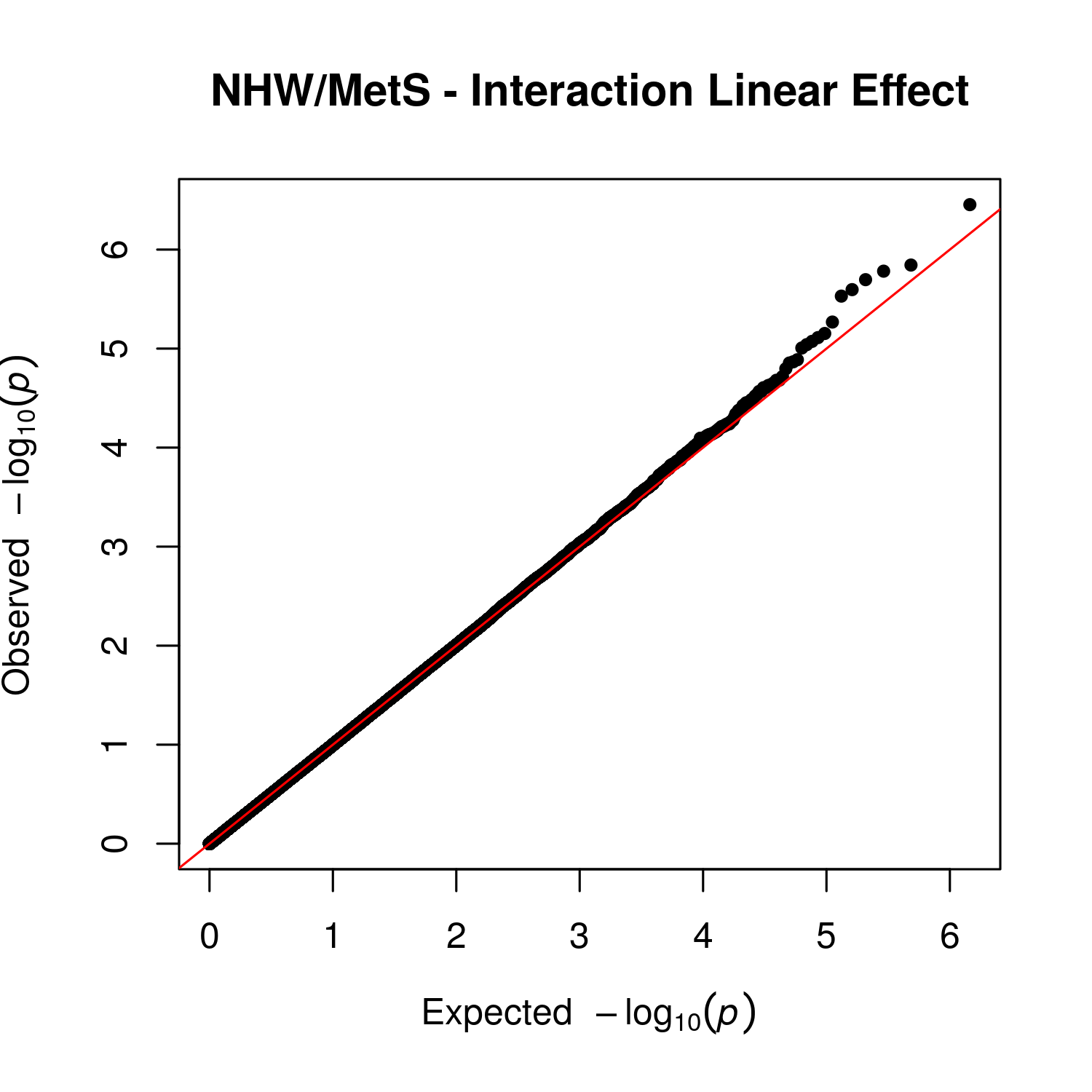

Supplemental Figure 9 – QQ Plot for Linear Interaction Effect between Cognitive Impairment and MetS Score in Non-Hispanic White, Metabolic Syndrome Group

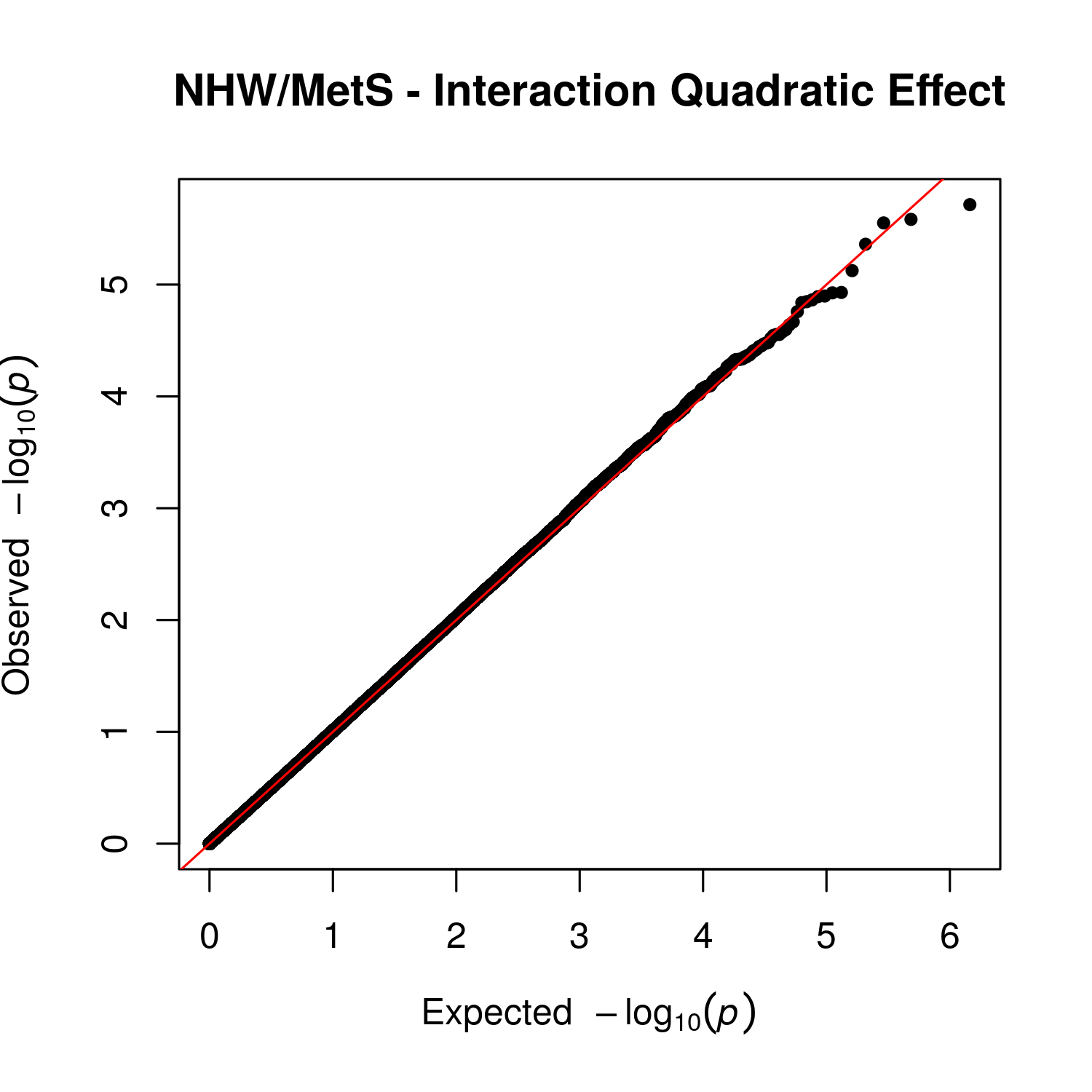

Supplemental Figure 10 – QQ Plot for Quadratic Interaction Effect between Cognitive Impairment and MetS Score in Non-Hispanic White, Metabolic Syndrome Group

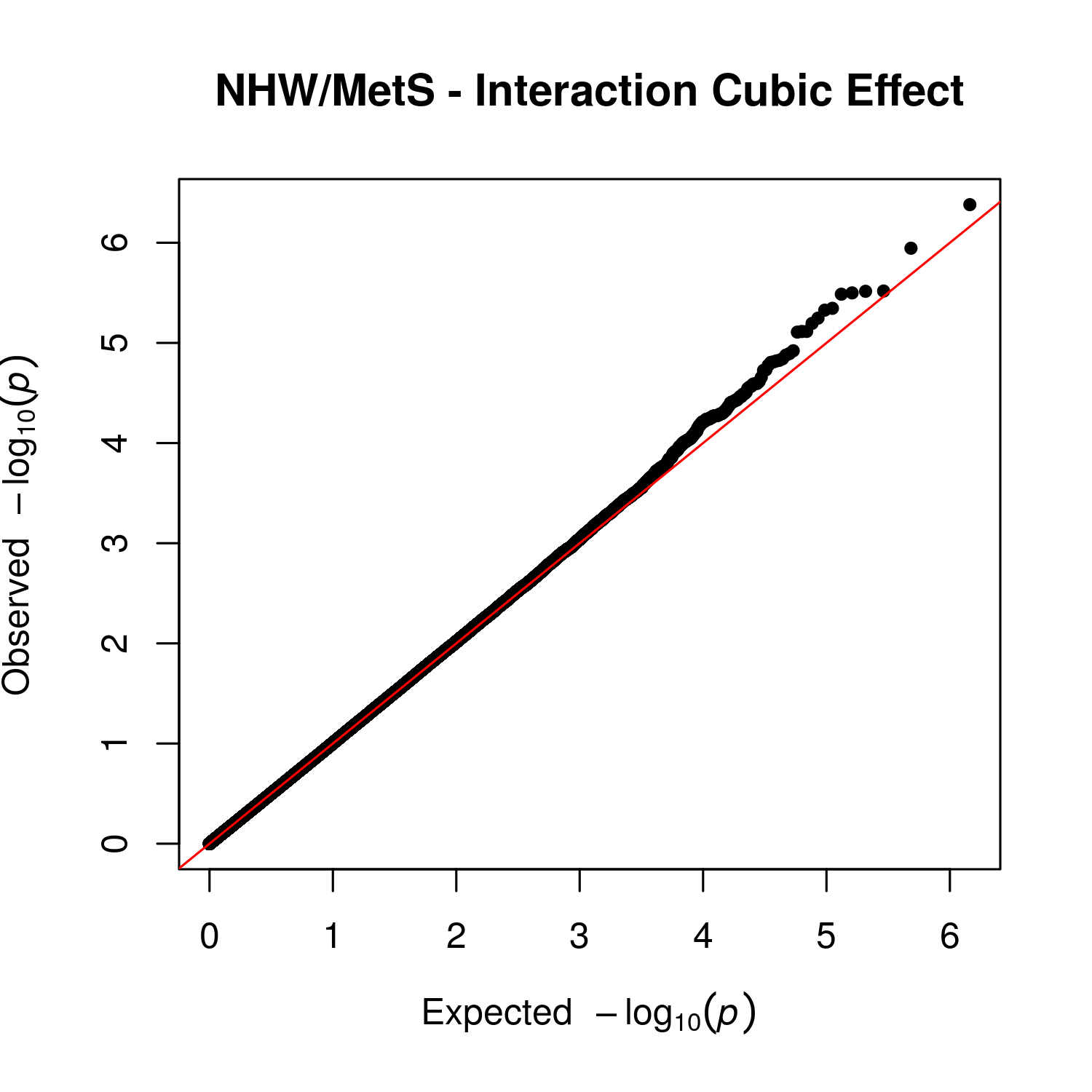

Supplemental Figure 11 – QQ Plot for Cubic Interaction Effect between Cognitive Impairment and MetS Score in Non-Hispanic White, Metabolic Syndrome Group

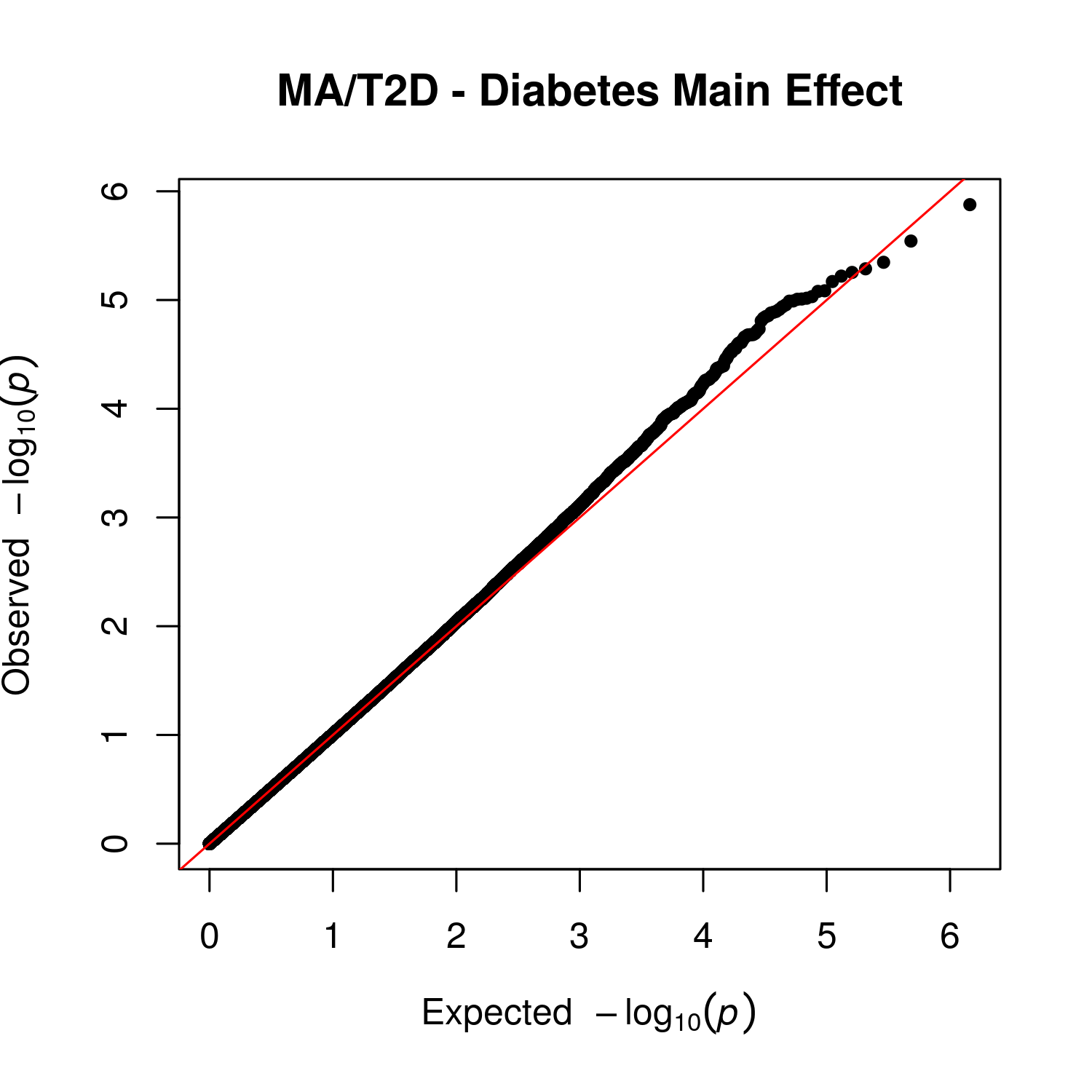

Supplemental Figure 12 – QQ Plot for Main Effect of Diabetes in Mexican American, Type 2 Diabetes Group

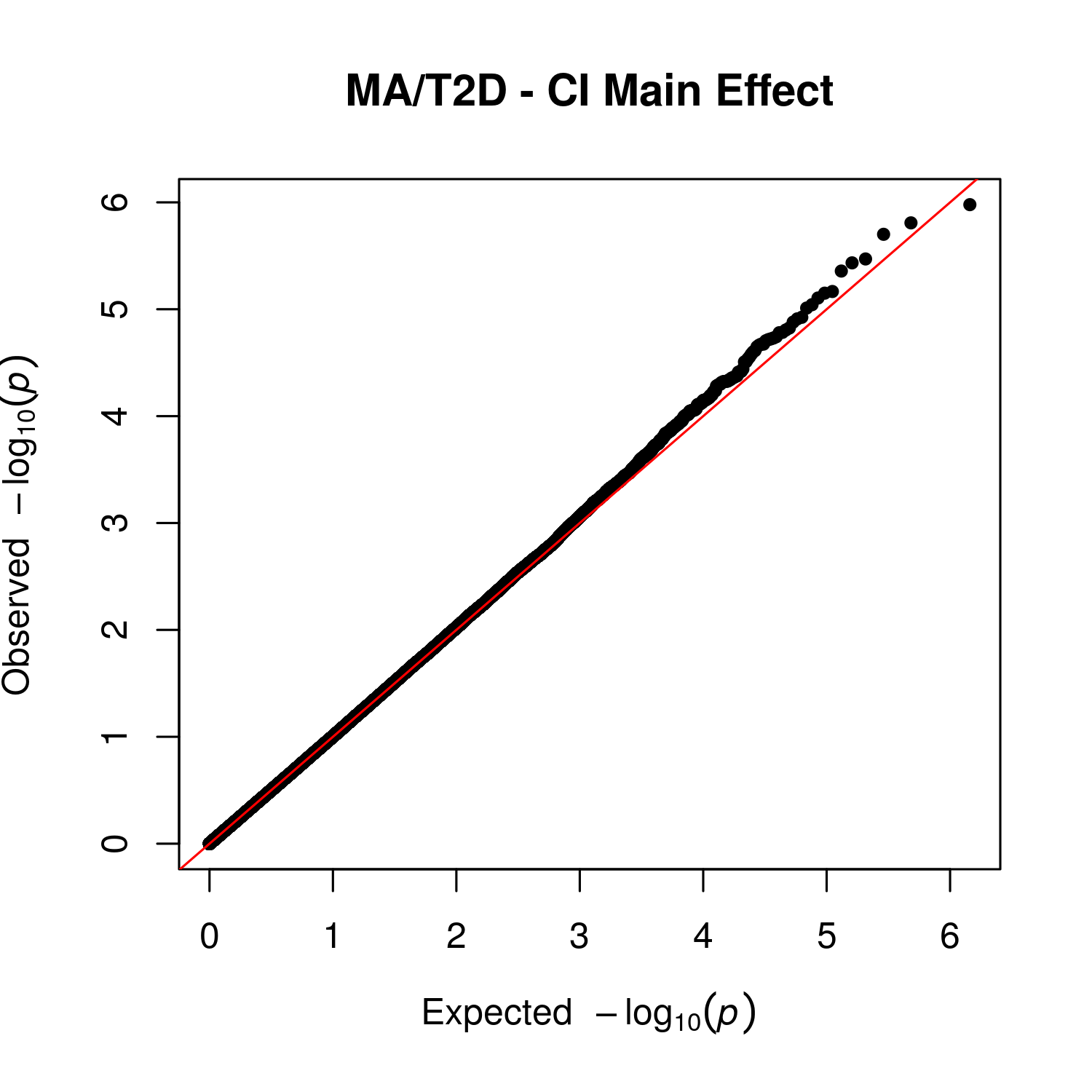

Supplemental Figure 13 – QQ Plot for Main Effect of Cognitive Impairment in Mexican American, Type 2 Diabetes Group

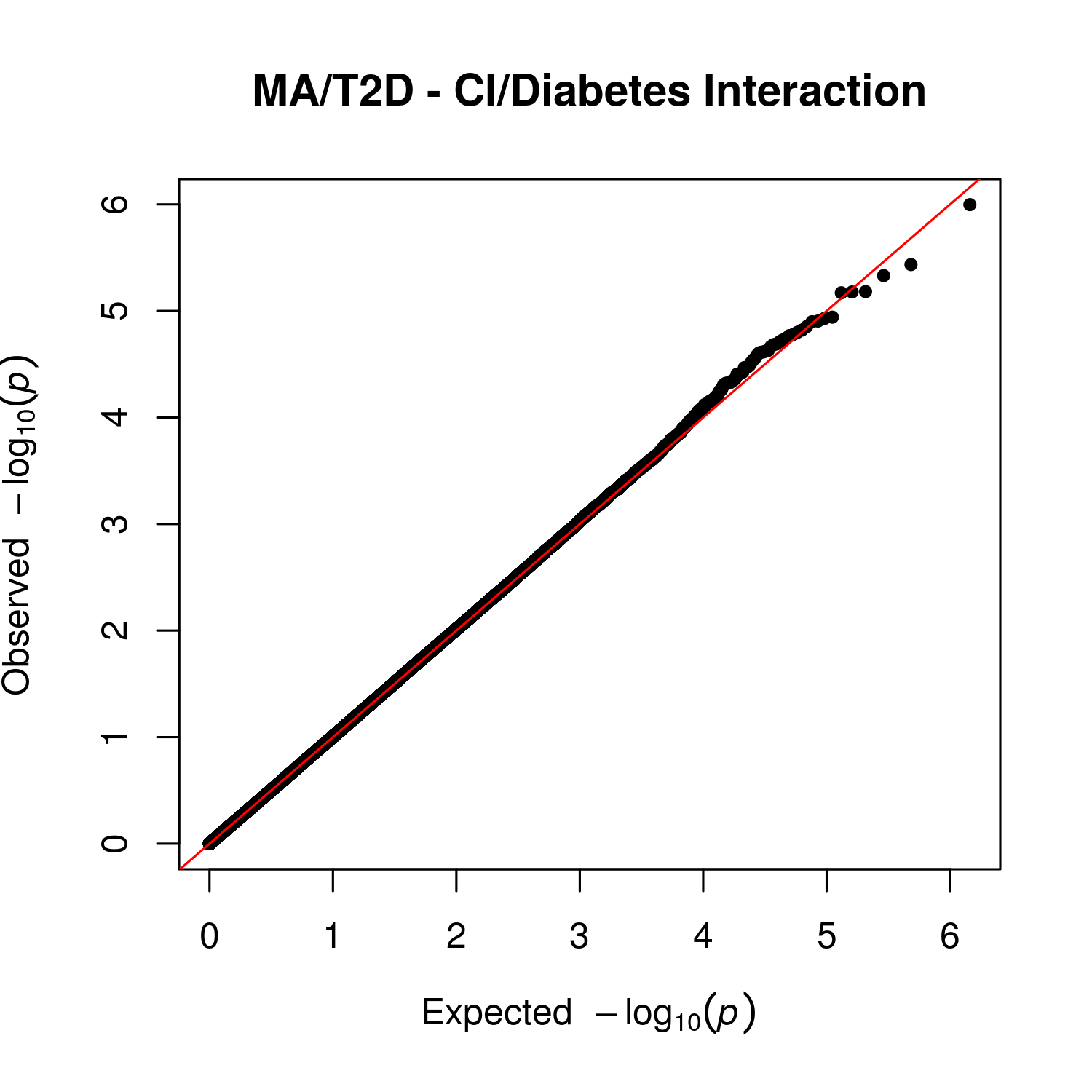

Supplemental Figure 14 – QQ Plot for Interaction Effect between Cognitive Impairment and Diabetes in Mexican American, Type 2 Diabetes Group

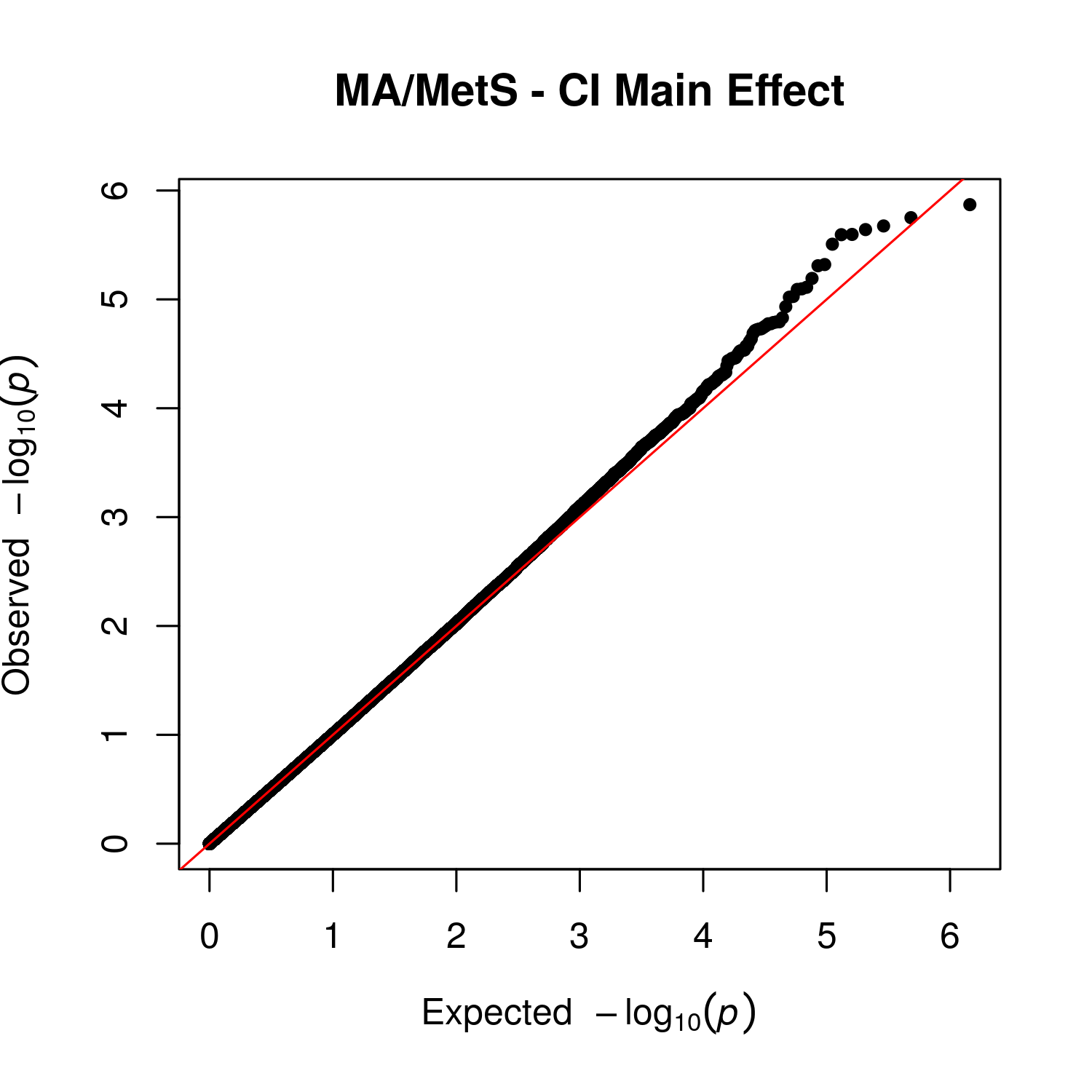

Supplemental Figure 15 – QQ Plot for Main Effect of Cognitive Impairment in Mexican American, Metabolic Syndrome Group

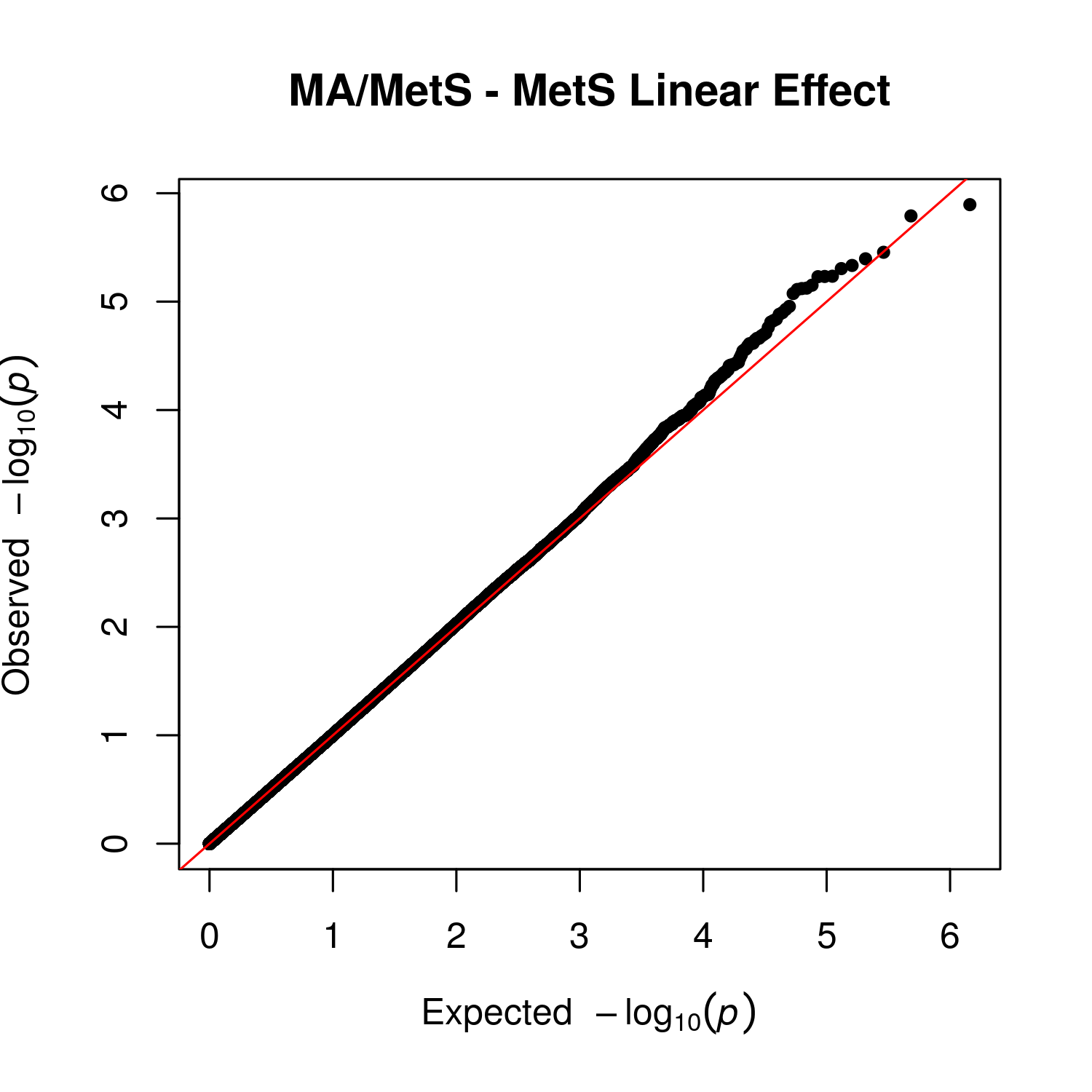

Supplemental Figure 16 – QQ Plot for Linear Main Effect of Metabolic Syndrome in Mexican American, Metabolic Syndrome Group

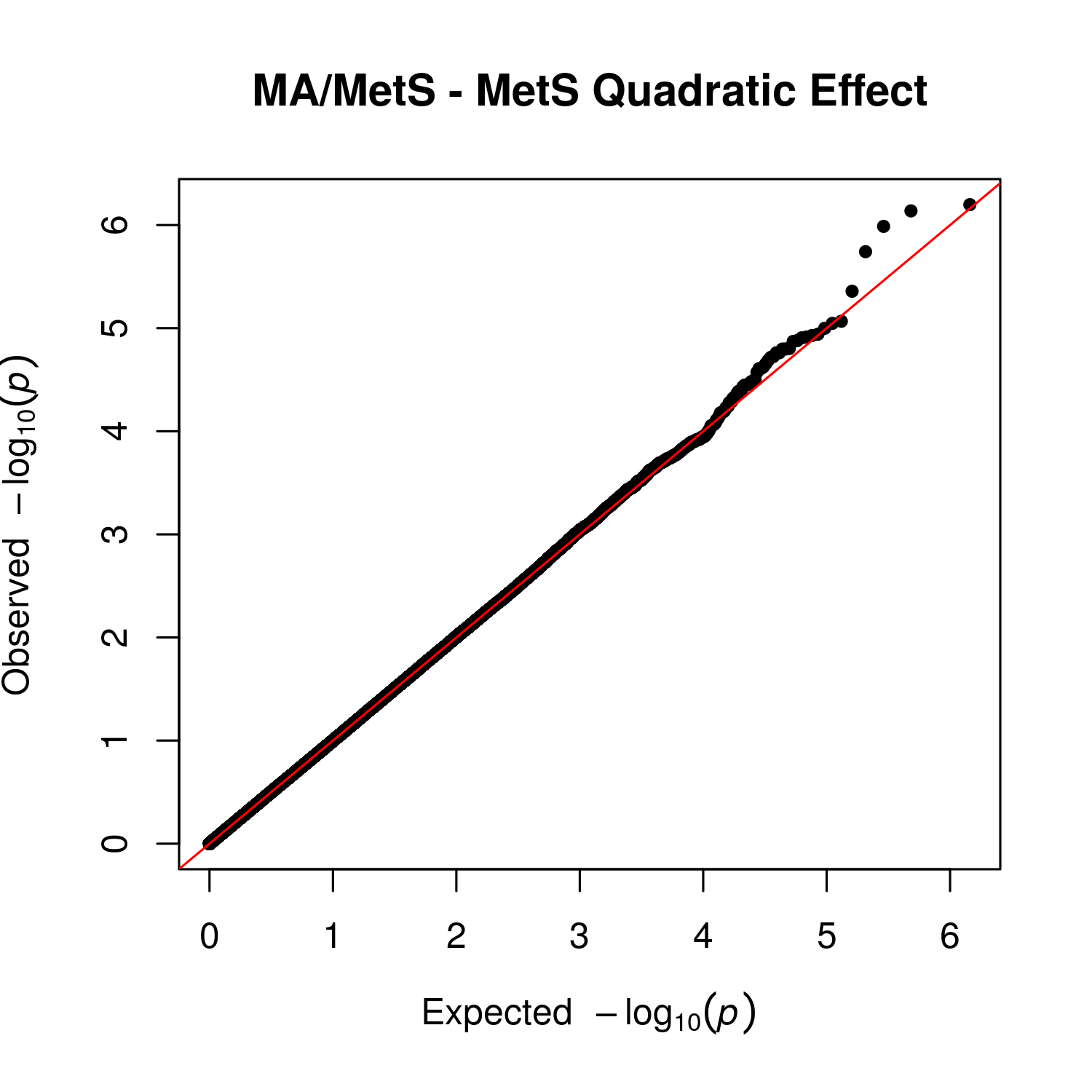

Supplemental Figure 17 – QQ Plot for Quadratic Main Effect of Metabolic Syndrome in Mexican American, Metabolic Syndrome Group

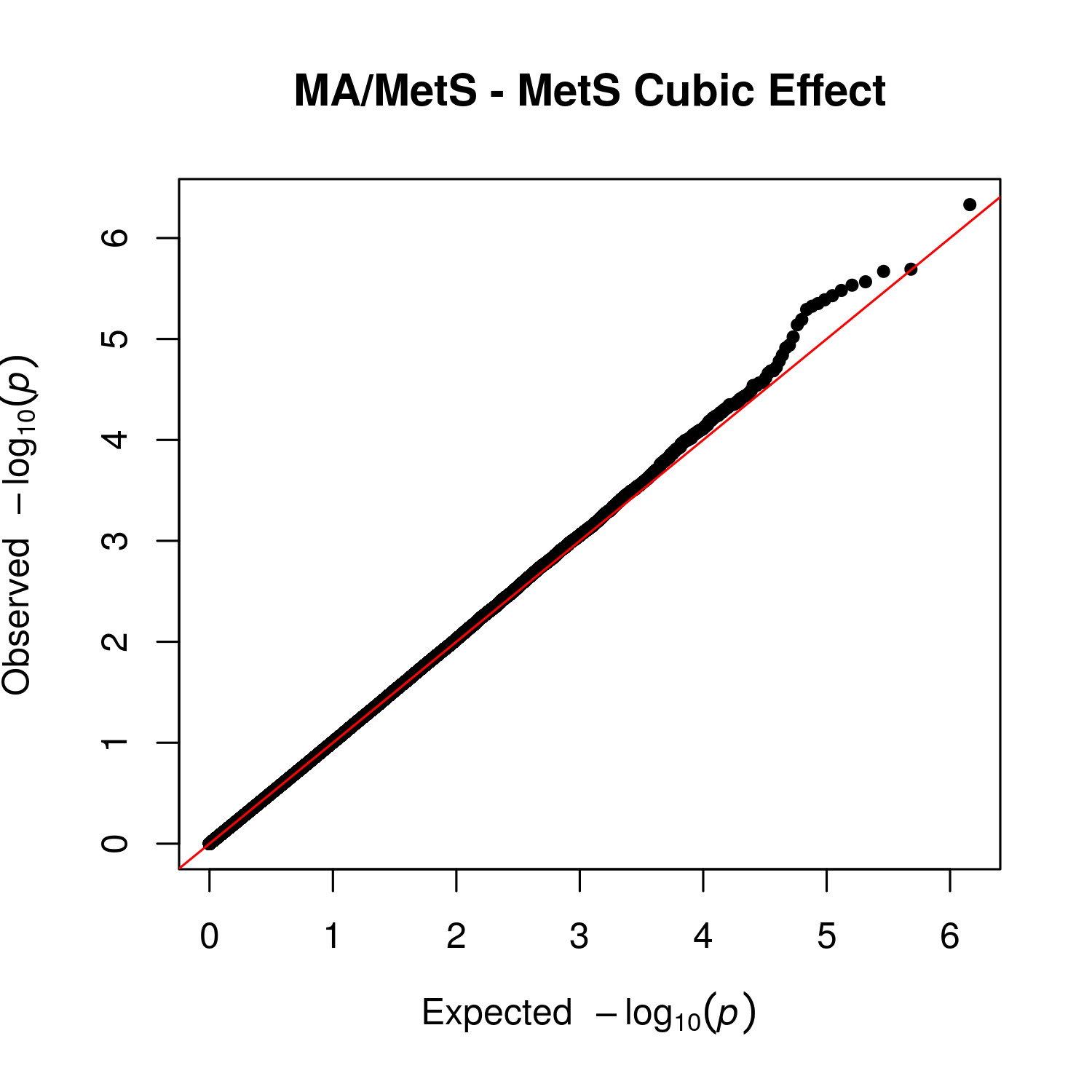

Supplemental Figure 18 – QQ Plot for Cubic Main Effect of Metabolic Syndrome in Mexican American, Metabolic Syndrome Group

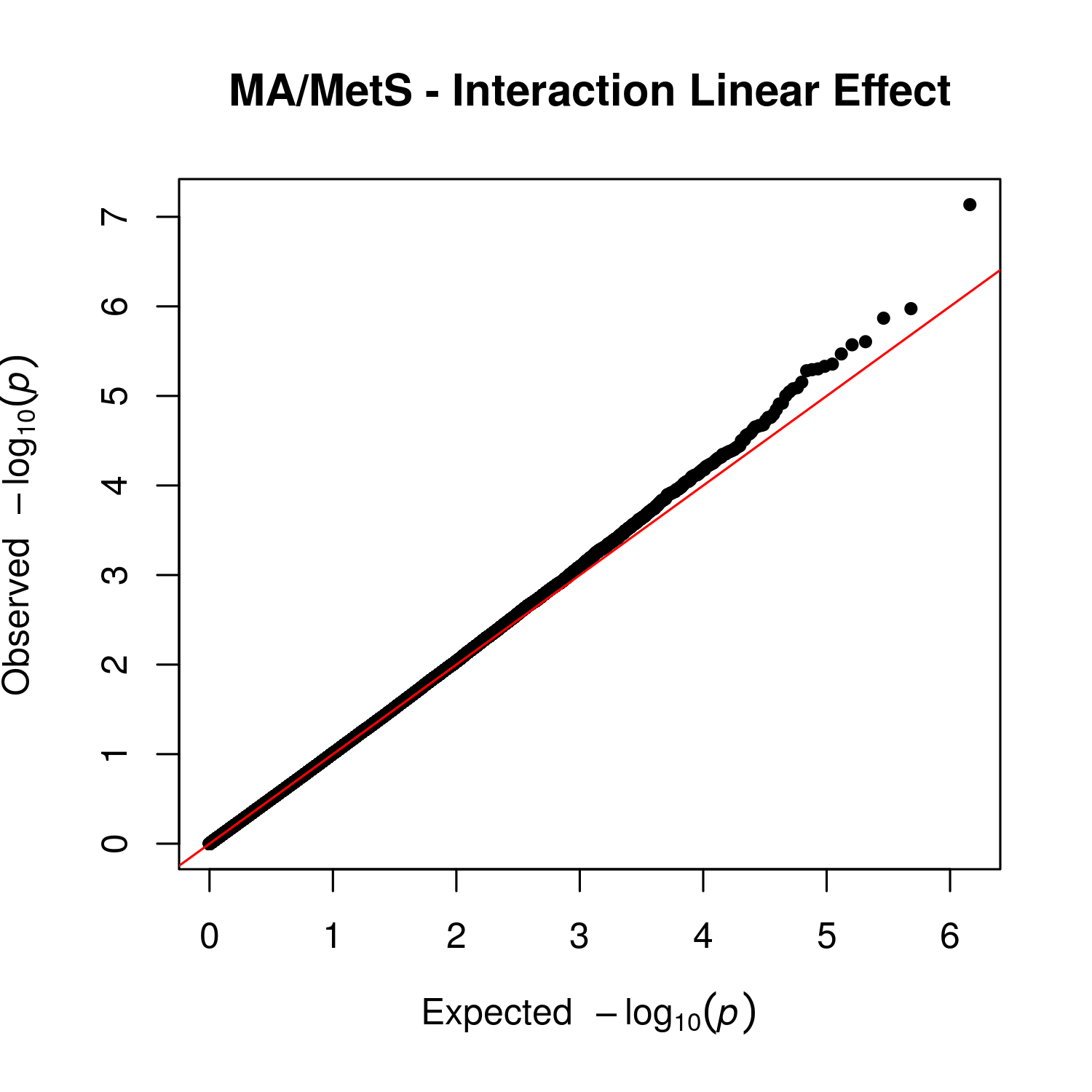

Supplemental Figure 19 – QQ Plot for Linear Interaction Effect between Cognitive Impairment and Metabolic Syndrome in Mexican American, Metabolic Syndrome Group

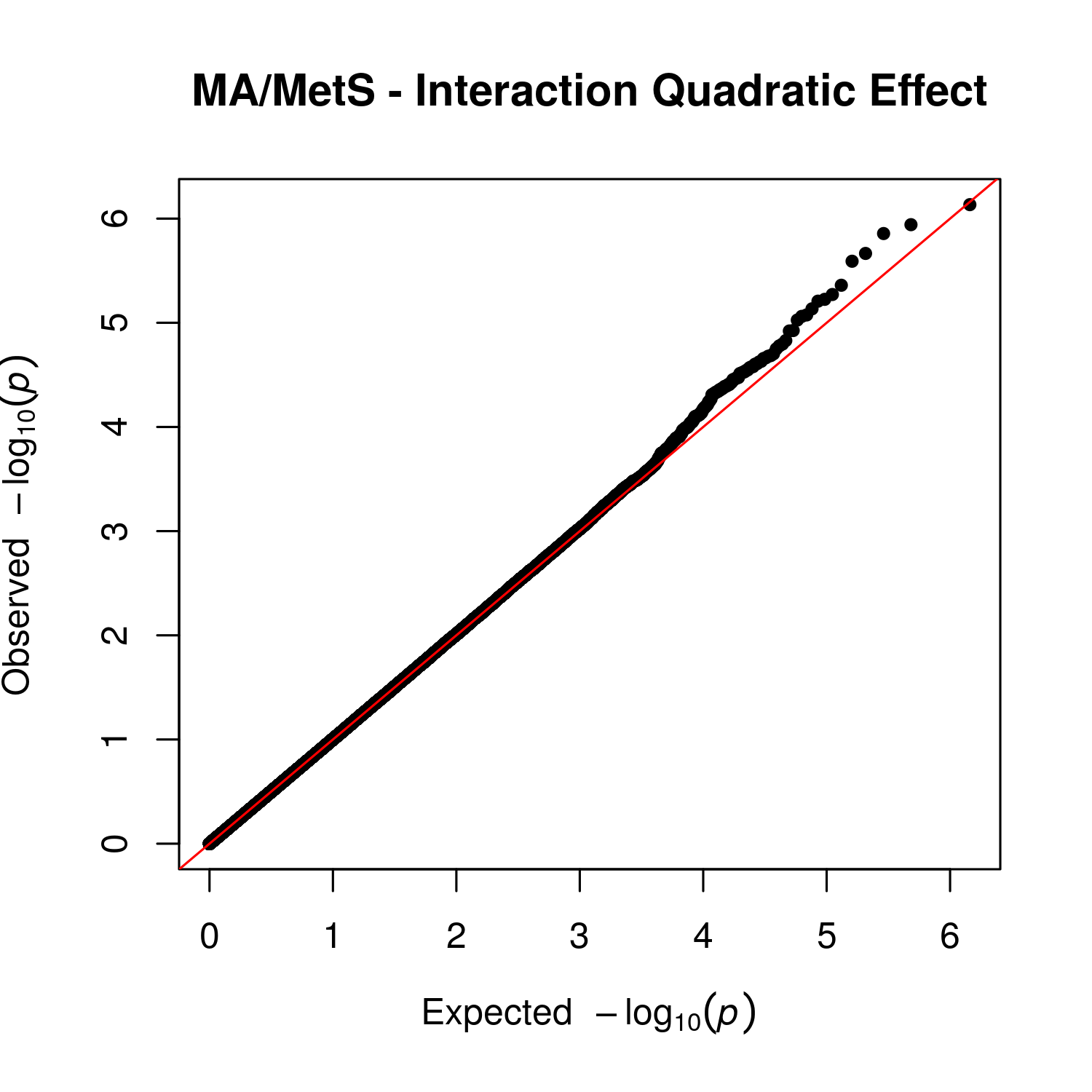

Supplemental Figure 20 – QQ Plot for Quadratic Interaction Effect between Cognitive Impairment and Metabolic Syndrome in Mexican American, Metabolic Syndrome Group
